# Diagnostic accuracy of the Panbio™ COVID-19 Antigen rapid test device for SARS-CoV-2 detection in Kenya, 2021: A field evaluation

**DOI:** 10.1101/2022.05.23.22275439

**Authors:** Irungu Karuga, Peninah Munyua, Caroline Ochieng, Bonventure Juma, Patrick Amoth, Francis Kuria, John Kiiru, Lyndah Makayotto, Ahmed Abade, Marc Bulterys, Elizabeth Hunsperger, Gideon O. Emukule, Clayton Onyango, Taraz Samandari, Beth A. Tippett Barr, Victor Akelo, Herman Weyenga, Patrick K Munywoki, Godfrey Bigogo, Nancy A. Otieno, Jackton Azenga Kisivuli, Edwin Ochieng, Rufus Nyaga, Noah Hull, Amy Herman-Roloff, Rashid Aman

## Abstract

**Background:** Accurate and timely diagnosis is essential in limiting the spread of severe acute respiratory syndrome coronavirus 2 (SARS-CoV-2) infection. Real-time reverse transcription-polymerase chain reaction (rRT-PCR), the reference standard, requires specialized laboratories, costly reagents, and a long turnaround time. Antigen rapid diagnostic tests (Ag RDTs) provide a feasible alternative to rRT-PCR since they are quick, relatively inexpensive, and do not require a laboratory. The WHO requires that Ag RDTs have a sensitivity ≥80% and specificity ≥97%.

**Methods:** This evaluation was conducted at 11 health facilities in Kenya between March and July 2021. We enrolled persons of any age with respiratory symptoms and asymptomatic contacts of confirmed COVID-19 cases. We collected demographic and clinical information and two nasopharyngeal specimens from each participant for Ag RDT testing and rRT-PCR. We calculated the diagnostic performance of the Panbio™ Ag RDT against the US Centers for Disease Control and Prevention’s (CDC) rRT-PCR test.

**Results:** We evaluated the Ag RDT in 2,245 individuals where 551 (24.5%, 95% CI: 22.8-26.3%) tested positive by rRT-PCR. Overall sensitivity of the Ag RDT was 46.6% (95% CI: 42.4-50.9%), specificity 98.5% (95% CI: 97.8-99.0%), PPV 90.8% (95% CI: 86.8-93.9%) and NPV 85.0% (95% CI: 83.4-86.6%). Among symptomatic individuals, sensitivity was 60.6% (95% CI: 54.3-66.7%) and specificity was 98.1% (95% CI: 96.7-99.0%). Among asymptomatic individuals, sensitivity was 34.7% (95% CI 29.3-40.4%) and specificity was 98.7% (95% CI: 97.8-99.3%). In persons with onset of symptoms <5 days (594/876, 67.8%), sensitivity was 67.1% (95% CI: 59.2-74.3%), and 53.3% (95% CI: 40.0-66.3%) among those with onset of symptoms >7 days (157/876, 17.9%). The highest sensitivity was 87.0% (95% CI: 80.9-91.8%) in symptomatic individuals with cycle threshold (Ct) values ≤30.

**Conclusion:** The overall sensitivity and NPV of the Panbio™ Ag RDT were much lower than expected. The specificity of the Ag RDT was high and satisfactory; therefore, a positive result may not require confirmation by rRT-PCR. The kit may be useful as a rapid screening tool for only symptomatic patients in high-risk settings with limited access to RT-PCR. A negative result should be interpreted based on clinical and epidemiological information and may require retesting by rRT-PCR.

## Introduction

The COVID-19 pandemic has had major negative socioeconomic effects. Prompt diagnosis of SARS-CoV-2 infection is critical in controlling the pandemic. Real-time reverse transcription-polymerase chain reaction (rRT-PCR) remains the reference standard for the diagnosis of SARS-CoV-2 infection. However, rRT-PCR has limitations, including the need for specialized laboratory, equipment, and staff, longer turnaround time (TAT) for clinical decision making and prevention measures, high cost, and erratic supply of reagents to conduct SARS-CoV-2 rRT-PCR due to increased demand (1,2). These limitations have increased the demand for more rapid, cheaper, and easy to perform testing methods.

Antigen rapid diagnostic tests (Ag RDTs) increase patients’ access to diagnosis of SARS-CoV-2 infection. The World Health Organization (WHO) recommends Ag RDTs with a minimum sensitivity and specificity ≥80% and ≥97%, respectively where rRT-PCR is unavailable or is associated with long TAT (3).

We evaluated the performance of the Panbio™ Ag RDT compared to the reference standard rRT-PCR. The Panbio™ Ag RDT is authorized in Kenya and received an emergency use license (EUL) from WHO (4). We also assessed the operational characteristics of the Panbio™ Ag RDT in the field setting.

## Methods

We conducted a prospective cross-sectional evaluation of the Panbio™ Ag RDT in 11 sites in Kenya between March and July 2021. The sites were located in Nairobi County (Nairobi remand Prison, Langata Women’s Prison, and Tabitha level 2 Clinic), Kiambu County (Kihara Sub-county Hospital), Nakuru County (Nakuru Prison), Nyeri County (Nyeri level 5 Hospital and Nyeri Prison), Kisumu County (Kisumu Prison-Kodiaga), Mombasa County (Coast General Teaching and Referral Hospital) and Siaya County (Siaya County Referral Hospital and St. Elizabeth Mission Hospital). The protocol was reviewed and approved by the Kenya Medical Research Institute’s (KEMRI) scientific ethical review unit (SERU). The US Centers for Disease Control and Prevention (CDC) institutional review board provided non-research determination approval. Further administrative approvals were obtained from the Ministry of Health Kenya, the National Commission for Science, Technology, and Innovation (NACOSTI) Kenya, the Kenya Prisons Service, and the county governments of Nakuru, Nyeri, Mombasa, Kiambu, and Siaya. Written informed consent was obtained for all enrolled participants. Written parental informed consent and assent were obtained for participants aged <18 years.

### Clinical case definition for enrollment

We enrolled persons of any age who presented to our study sites with respiratory symptoms (symptomatic participants) that met any of three case definitions, including acute respiratory infection (ARI,) defined as cough or difficulty in breathing or sore throat or coryza, with the onset of symptoms <2 weeks; severe acute respiratory infection (SARI) defined as fever or temperature ≥38 °C and cough, with onset of symptoms <10 days and requiring hospitalization; or influenza-like illness (ILI) defined as temperature ≥38°C and cough, with onset of symptoms <10 days. Body temperature measurement was done using infrared thermometers placed 3 to 5 centimeters from the temple. We also enrolled asymptomatic close contacts of individuals with confirmed SARS-CoV-2 infection exposed between two days before and 14 days after symptoms onset or confirmation of infection. Individuals exposed to persons with confirmed SARS-CoV-2 infection in the following setups were considered close contacts in the following scenarios: working in close proximity or sharing the same environment; face-to-face contact within 1 meter for more than 15 minutes; traveling together in any kind of conveyance; living in the same household; healthcare-associated exposure, including providing direct care for COVID-19 patients, visiting patients or staying in the same close environment.

### Data and specimen collection

Individuals who met the case definition and consented were enrolled and assigned unique identifiers. We collected demographic information, and clinical information via electronic questionnaires on smartphones. Data was stored in a secured cloud server. Clinical information collected included the range and duration of respiratory symptoms, presence of comorbidities, and reported history of exposure to confirmed COVID-19. Two nasopharyngeal swabs were collected by a surveillance officer [nurse, clinical officer, or laboratory technologist] trained to collect respiratory specimens from each of the nostrils a few seconds apart. The first specimen was collected using the swab provided in the Panbio™ Ag RDT kit. The second specimen was collected using a polyester-tipped aluminum shafted swab and immediately placed in viral transport media at +2 to +8^0^C and shipped to the CDC-supported KEMRI laboratories in Nairobi or Kisumu for rRT-PCR testing where long-term storage was done at −80°C.

### Rapid SARS-CoV-2 antigen testing

Rapid testing was conducted using the Panbio™ Ag RDT (Abbott Rapid Diagnostics, US. Ref. 41FK10) per manufacturer instructions by surveillance officers. The Panbio™ Ag RDT is a lateral flow immunoassay that detects the nucleocapsid (N) protein in nasal and nasopharyngeal specimens(5). Results were read and documented independently by two surveillance officers within 15 to 20 minutes. Positive, negative, and invalid results were interpreted according to the manufacturer’s recommendations. Each reader documented the test results in the electronic questionnaire independently. In the event the two readers documented different results, the final result was indicated as indeterminate.

### Assessing the operational characteristics of the Panbio™ Ag RDT kit

We assessed operational characteristics of the Panbio™ Ag RDT in the field setting including clarity of kit instructions, technical complexity or ease of use, and the ease of interpretation of results via a standardized questionnaire administered to users on site.

### RT-PCR testing in the Laboratory

According to the manufacturer’s instructions, total nucleic acid material was extracted from 200µL of the nasopharyngeal specimen using either the Ambion MagMax Total RNA isolation kit (Thermo Fisher Scientific, Greenville, North Carolina, USA) performed on a semi-automated KingFisher flex machine (Thermo Fisher Scientific) or Standard M Spin-X Viral RNA Extraction Kit (SD biosensor) according to the manufacturer’s instructions. We then eluted 60uL of total RNA from the extracted sample. The SARS-CoV-2 rRT-PCR was performed using the TaqPath 1-step Multiplex rRT-PCR master mix (Thermo Fisher Scientific) or using the GoTaq® Probe 1-Step RT-qPCR System (Promega A6120). Both systems are compatible with the rRT-PCR CDC IDT kit. Each of the 15µL reaction volumes contained 5µL of master mix, 8.5µL of nuclease-free water, and 1.5µL of CDC-IDT primer/probe mix for each of the N1, N2, and RNP3 genes separately. The eluate was then amplified using QuantStudio™ 5 Real-Time PCR System, 0.1ml block (Thermo Fisher Scientific), and analyses were done using QuantStudio 3 and 5 Real-Time PCR System Software version 1.5.1 (Thermo Fisher Scientific). SARS-CoV-2 assay targets, N1 and N2 (N1 and N2) were run simultaneously with the human ribonuclease P gene (RNP) control to monitor the quality of the nucleic acid extraction, specimen quality, and presence of reaction inhibitors for assay performance. The positive, negative, and human specimen controls were included in all assays.

### RT-PCR testing

When all controls exhibited expected performance, a positive result for SARS-CoV-2 was considered, if all assay amplification curves crossed the threshold line within Ct <40. If SARS-CoV-2 (N1 and N2) assay results were negative, then the test result was reported as negative. If only one target (N1 and N2) were positive, then it was designated as inconclusive, and retesting was conducted. If upon repeating the test, the results remain inconclusive, then the final result was reported as inconclusive. Results were relayed back to the specific sites for patient management.

### Sample size calculation

To evaluate the diagnostic performance of the Panbio™ Ag RDT device, we assumed a prevalence of 10% among symptomatic individuals and 5% in asymptomatic contacts of laboratory-confirmed persons based on current SARS CoV 2 local surveillance data. We assumed a sensitivity of 90% for the Panbio™ Ag RDT with 7.5% margin of error, and after adjusting by 10% for other concerns we anticipated enrolling 770 symptomatic patients and 1,540 close contacts of confirmed cases.

### Statistical analysis

Statistical analysis was conducted using Stata version 15.1 (College Station, Texas). Measures of central tendency and dispersion were calculated for continuous variables such as age, days since onset of symptoms, temperature, and cycle threshold (Ct) values. We calculated frequencies and proportions for categorical variables such as age group, occupation, and Ct value cut-offs. The 95% confidence intervals for categorical variables were calculated using the normal approximation method (Wald’s test). We compared proportions between the symptomatic and asymptomatic groups by calculating their Z scores. P-values <0.05 were considered statistically significant.

We calculated the sensitivity, specificity, and positive and negative predictive values for the Panbio™ Ag RDT against rRT-PCR as the reference standard. Stratified analysis of performance was done by age, day’s post-onset of symptoms, and Ct values. The measure of agreement between Ag RDT and rRT-PCR results was evaluated using Cohen’s Kappa statistic. We interpreted the strength of agreement as slight agreement (κ, 0-0.2), fair (κ, 0.21-0.4), moderate (κ, 0.41-0.6), substantial (κ, 0.61-0.8), and almost perfect (κ, 0.81-1.0) as described by Landis *et al*. (6).

## Results

### Enrolled participant characteristics

We enrolled 2,279 participants, where 38.9% had at least one respiratory symptom. The median age was 31.0 years, and over half (60.9%) were male (Table 1). The median duration post-onset of symptoms among symptomatic participants was three days, with the majority (67.8%) presenting within 5 days.

**Table 1:**
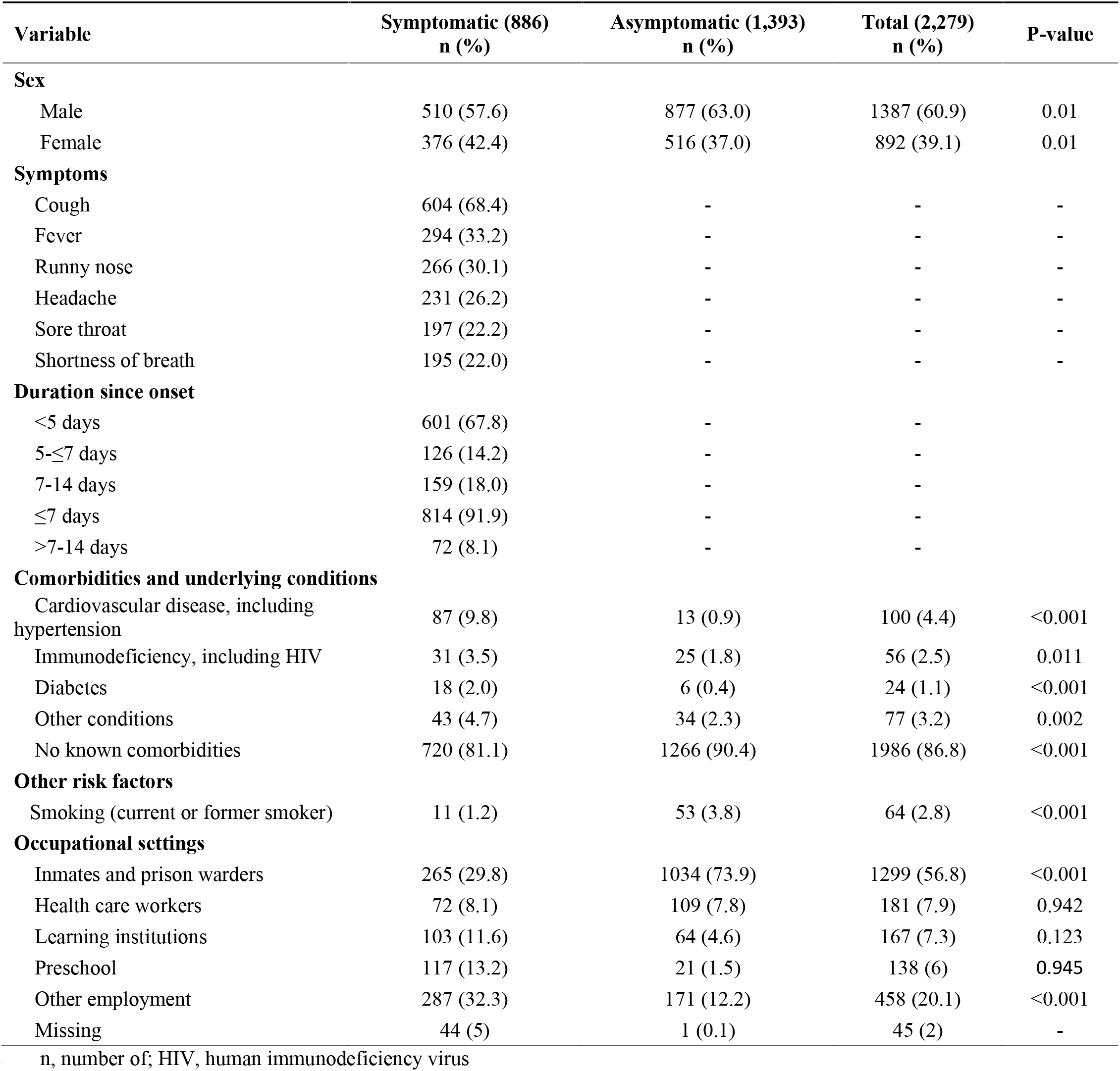
Clinical and demographic characteristics of participants in the field evaluation of the Panbio™ Ag RDT in Kenya, 2021

#### Laboratory findings

##### Panbio™ Ag RDT test results

Among the 2,277 tested using the Panbio™ Ag RDT device, 12.4% were positive (Fig 1). There were no invalid or indeterminate Ag RDT results. The positivity was significantly higher among symptomatic individuals (18.7%) compared with the asymptomatic (8.4%,) individuals.

**Fig 1:**
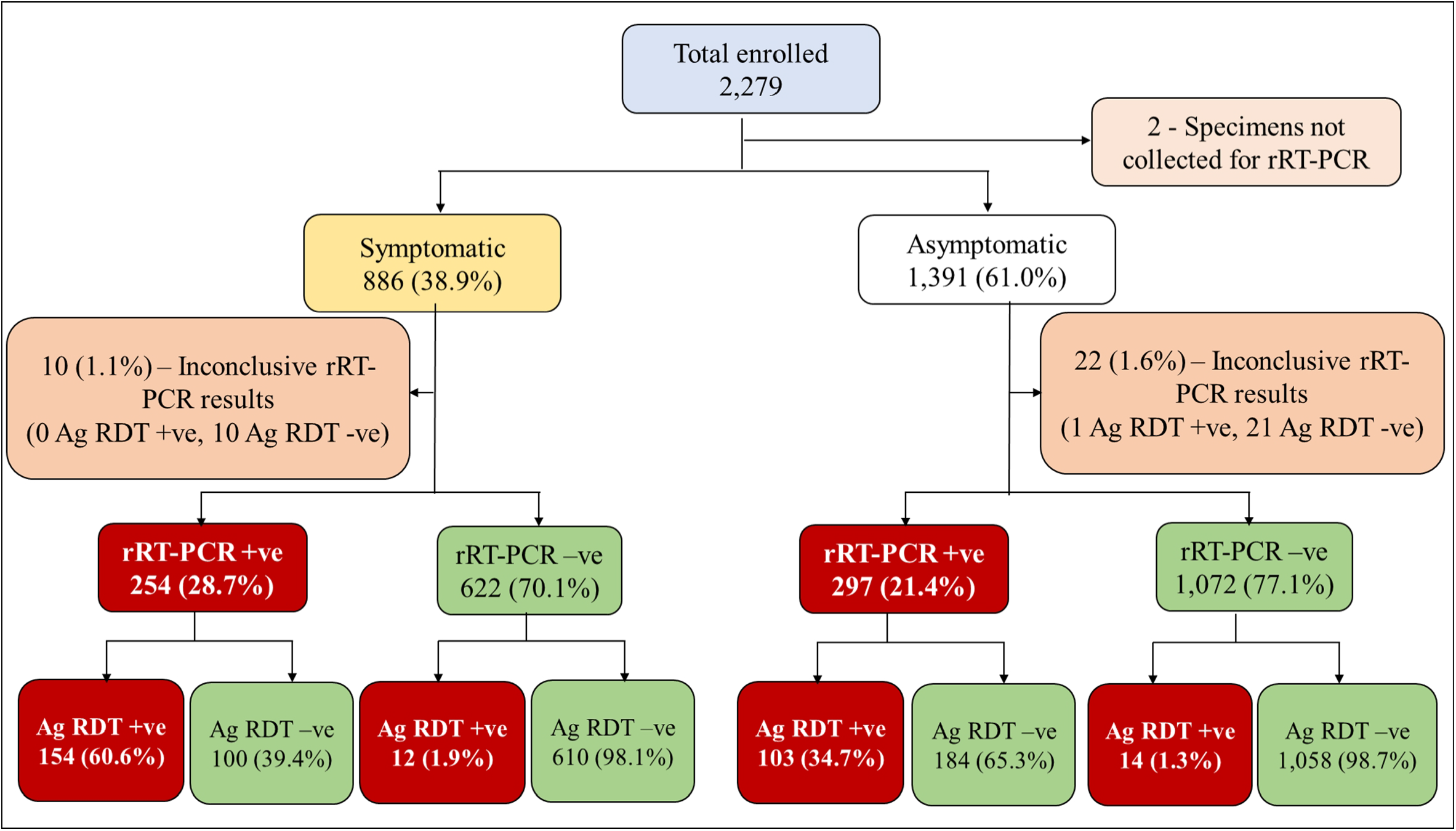
Flow diagram of enrolment of study participants and distribution by clinical status and test results in the field evaluation of the Panbio™ Ag RDT in Kenya, 2021 Ag RDT, Antigen rapid diagnostic test; rRT-PCR, real-time reverse transcriptase-polymerase chain reaction

**Fig 2:**
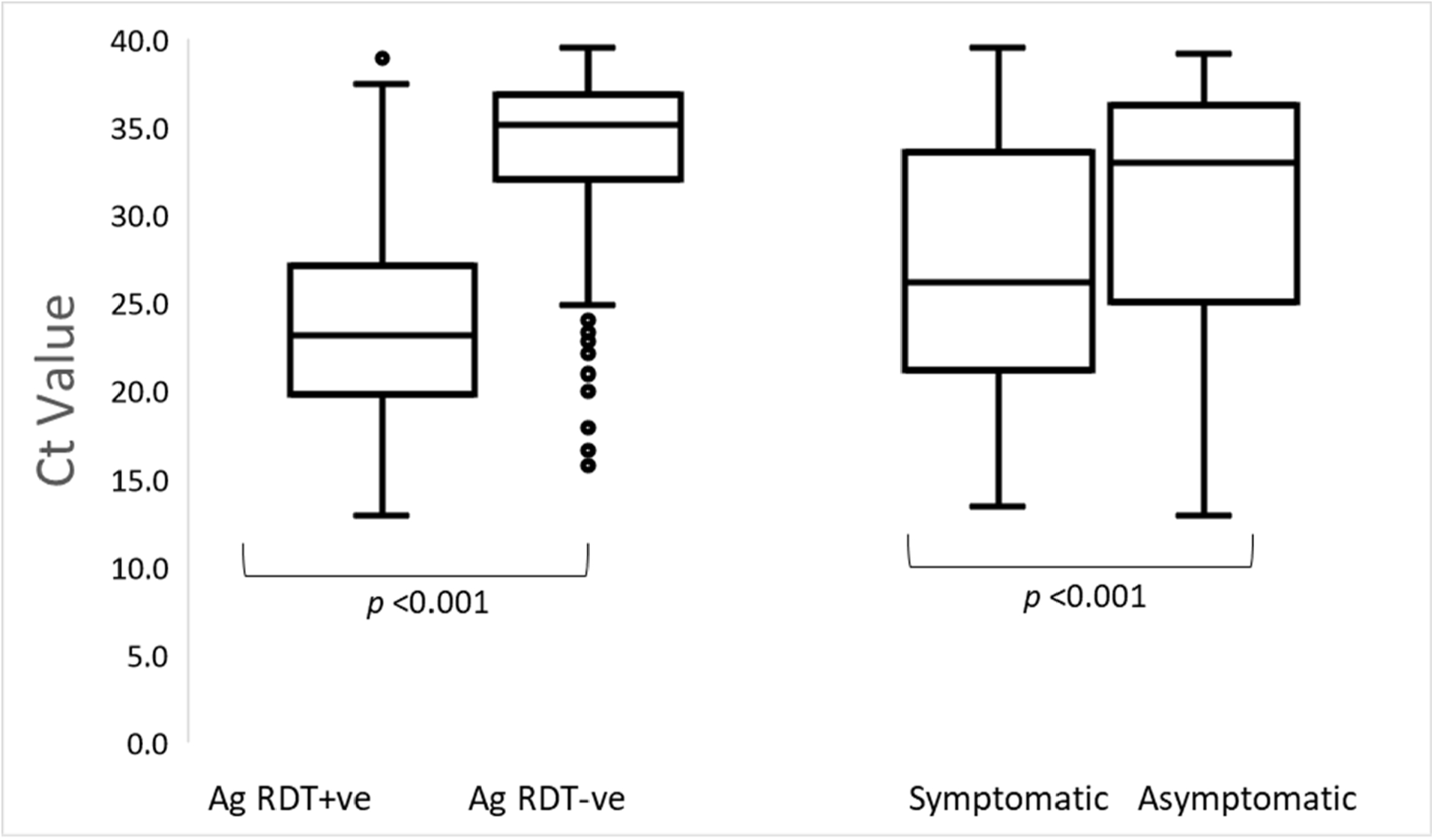
Distribution of Cycle threshold (Ct) values for rRT-PCR positive samples by Ag RDT result and by presence/absence of symptoms, Kenya 2021 (n=551) Ag RDT, Antigen rapid diagnostic test; Ct, cycle threshold

##### rRT PCR test results

We tested 2,277 specimens by rRT-PCR, 24.2% tested positive and 1.4% were inconclusive. Positivity (28.7%,) was higher among symptomatic participants compared to asymptomatic (21.4%) ones. Among the 32 inconclusive rRT-PCR results, 22 (68.8%) were from asymptomatic individuals, and 31 (96.9%) tested negative by Ag RDT (Fig 1). The overall median Ct value of the 551 rRT-PCR positive results was 30.2 (IQR 22.9-35.6). Symptomatic individuals had a significantly lower median Ct value of 26.2 compared to 33.0 among asymptomatic individuals (*p* <0.001). The median Ct value was significantly lower among those with Ag RDT positive results (23.2) compared to negative test results (35.1) (*p* <0.001) (**Error! Reference source not found**.).

***<u>Error! Reference source not found.</u>***

### Performance of Panbio™ Ag RDT

We reviewed 2,245 paired Ag RDT/rRT-PCR records for the performance analysis. The overall sensitivity of the Panbio™ Ag RDT was 46.6% and specificity was 98.5% (Table 2). At the observed SARS-CoV-2 prevalence (24.5%) the positive predictive value (PPV) was 90.8% and the negative predictive value (NPV) was 85.0% (Table 2; S1 Fig and S2 Fig). The overall agreement between the Ag RDT and rRT-PCR results was moderate (*k*=0.5,), substantial (*k=0*.*7*) among the symptomatic, and fair (*k=0*.*4*) among the asymptomatic. The performance of the Panbio™ Ag RDT is summarized in Table 2.

**Table 2:**
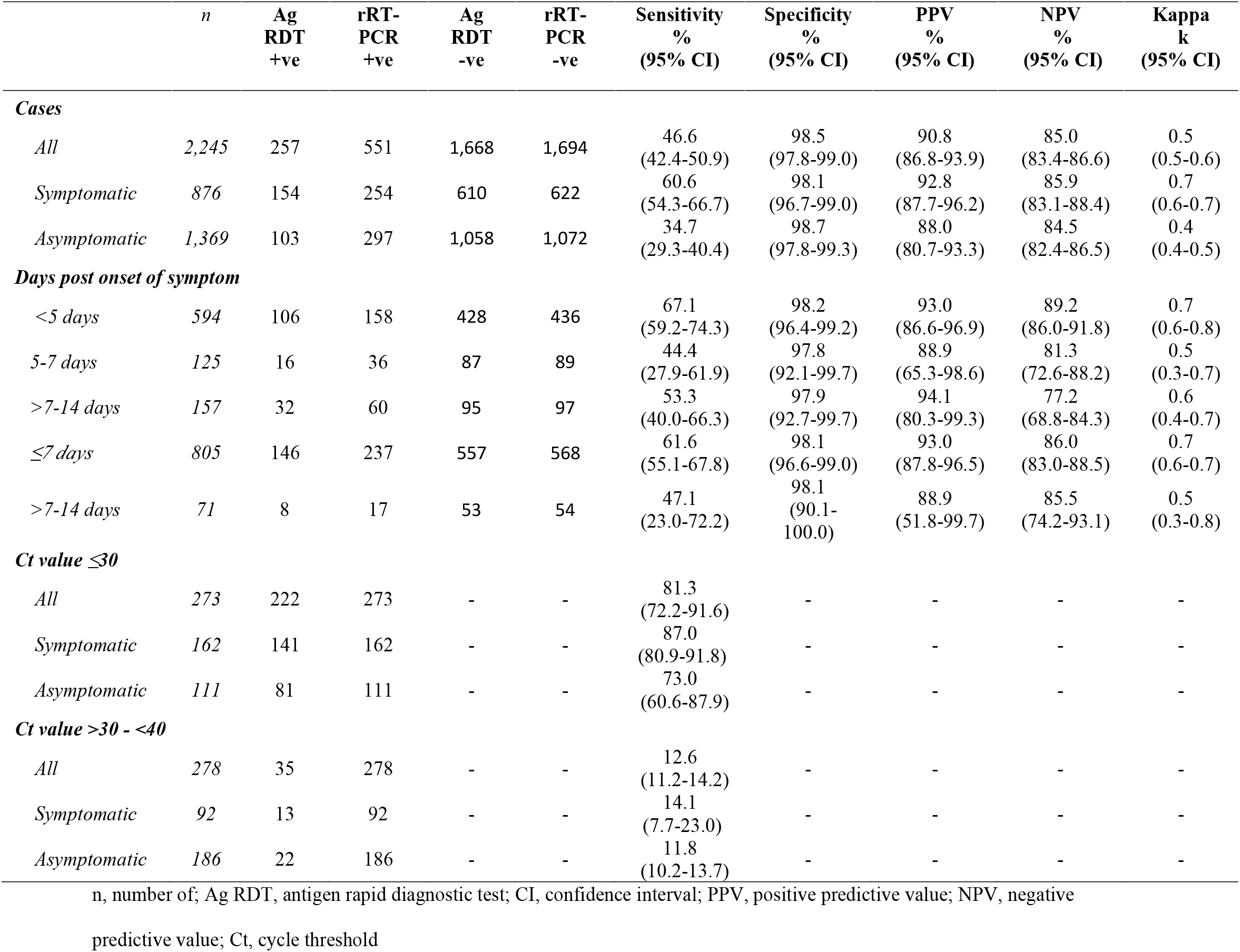
The performance of the Panbio™ Ag RDT by clinical status, and Ct value

Among the symptomatic participants who were positive for SARS-CoV-2 by rRT-PCR, 100 (39.4%) had a false negative result by Ag RDT. Two percent (12/632) who were negative for SARS-CoV-2 by RT-PCR had a false positive result by Ag RDT. Among the asymptomatic individuals, 65.3% (n=194) had a false negative Ag RDT result, and 1.3% (n=14) had a false positive result by Ag RDT.

Among the 876 symptomatic participants, the sensitivity was 60.6% (95% CI: 54.3-66.7%) (S3 Fig). Sensitivity in those with days post onset of symptoms <5 was 67.1% and declined to 53.3% among those with onset >7 days. Among the asymptomatic, sensitivity was significantly lower at 34.7% (95% CI: 29.3-40.4%) compared to symptomatic individuals (*p* <0.001). Specificity remained high in symptomatic (98.1%, 95% CI: 96.7-99.0%) and asymptomatic individuals (98.7%, 95% CI: 96.7-99.0%) (S4 Fig).

Regardless of the presence or absence of symptoms, the overall sensitivity of the Ag RDT test in 273 specimens with Ct values ≤ 30 was 81.3% (95% CI: 72.2-91.6). This reduced to 12.6% (95% CI: 11.2-14.2%) among the 278 individuals with Ct >30 (Table 2).

#### Evaluation of the user’s experience of the Panbio™ Ag RDT

A total of 17 surveillance officers comprising 10 (59%) clinicians and 7 (41%) laboratory staff completed the survey. All 17(100%) surveillance officers found the test procedure easy to perform, and the manufacturers’ instructions were found to be clear and easy to follow. Four (23.5%) of the surveillance officers noted the difficulty in dispensing a sample onto the test device from the extraction tube when nasopharyngeal specimens were collected from individuals with thick mucus.

## Discussion

We present findings of a field evaluation of the Panbio™ Ag RDT conducted among both symptomatic patients with onset of symptoms up to 14 days and asymptomatic individuals who had contact with individuals with confirmed SARS-CoV-2 infection within two days before detection of the index case and up to 14 days after exposure. Our evaluation was conducted in the third COVID-19 wave in Kenya(7,8).

The overall sensitivity was low (46.6%), detecting slightly less than half of the rRT-PCR confirmed cases, therefore, missing more than half and classifying them as not having SARS-CoV-2 infection when they were indeed infected. The overall sensitivity we report was lower than observed in studies that enrolled both symptomatic and asymptomatic individuals in a multicenter study in Spain conducted among individuals of any age (57.3%, 95%CI: 48.3-95.8%) (9), Spanish adults in primary care centers (71.4%, 95% CI: 63.1-78.7%) (10), among Spanish individuals of any age (73.3 %, 95 % CI: 62.2–83.8%) (11), and persons of any age presenting within 7 days of onset or exposure (90.5%, 95% CI 87.5-93.6%) (12).

The low overall sensitivity observed may be due to the high proportion (61.1%) of asymptomatic participants, and the higher Ct values we observed in these individuals. The lower performance could also be due to the relatively younger (median age 31 years) population we enrolled compared to other studies we reviewed where the median age was higher (39-51.5 years) (11,13). A recent study demonstrated increasing viral load with increasing age (14); higher sensitivities have been observed among individuals with higher viral loads compared to lower ones. The alpha variant and delta variants predominated our evaluation in Kenya (15,16). While some literature indicates that variants of concern (17,18) do not reduce the sensitivity and specificity of Ag RDTs, a recent study revealed that sensitivity of the Panbio™ Ag RDT declined significantly in individuals infected with the alpha variant (53.0%) compared to those with non-alpha variants (89.0%) even after adjusting for viral load (p <0.002) (19). Mutations occurring in the N protein of the Alpha variant may not be detected by the Panbio™ Ag RDT (20).

The sensitivity among symptomatic individuals (60.6%) was lower than the WHO recommendation of 80% (21). The sensitivity among symptomatic individuals with onset of symptoms within 7 days was also lower than that reported by the manufacturer on specimens tested on nasopharyngeal specimens (61.6% vs. 91.1%) (5). This is possibly due to the use of specimens with high viral loads by the manufacturer’s study for approval as hypothesized by *Albert et al*., *2021* in an evaluation where SARS-CoV-2 cultures were done (1). Similar sensitivity was observed in symptomatic Mexican adults (58.1%, 95% CI: 54.9-61.3%) (22). However, the sensitivity we observed among symptomatic individuals was higher compared to that reported in a multicenter evaluation conducted among symptomatic children (60.6% vs. 45.4%) with the onset of symptoms within five days (13).

In our evaluation, sensitivity was higher among symptomatic individuals compared to asymptomatic ones (60.6% vs. 34.7%). Similar findings were observed in other evaluations with higher sensitivity among symptomatic individuals compared to asymptomatic in a study in Mexico (58.1% vs. 26.3%) (22), in Spain (80.4% vs. 56.6%) (10), among Greek children (95.2% vs. 22.2%) (23), and among Swiss children (73% vs. 43%) (24). However, a German study found similar sensitivities between the two groups (86.8% vs. 85.7%) (25).

Individuals with Ct value ≤30 had higher sensitivity compared to those with Ct value >30, as observed in other evaluations of Panbio™ Ag RDT (11). The same has been observed in other evaluations considering Ct values ≤30 (25–27). We also observed higher viral loads among symptomatic individuals (median Ct = 26.2) compared to asymptomatic individuals (median Ct = 33.0) from our evaluation (p <0.001). Similar findings have also been reported recently (28,29). In contrast, some studies found no difference in viral load between the two groups (30,31,) while others found higher viral load among asymptomatic individuals (32,33). Higher sensitivity has been observed among individuals presenting in the early symptomatic phase of their infection (12,22,25–27,34–36). While rRT-PCR testing for SARS-CoV-2 is the reference standard for diagnosis limited access to molecular tests in our setting and provision of results in a clinically relevant timely manner to inform case management limits their clinical utility. Therefore, given reasonable sensitivity, individuals with a negative test in an individual meeting the clinical and epidemiological criteria may require a confirmatory test by RT-PCR. The variation in sensitivity by presence or absence of symptoms, symptom duration, and viral load indicates that the kit may only be useful among individuals with high viral loads, especially those with symptoms.

The PPV (90.8%) observed at the prevalence of SARS-CoV-2 infection of 24.5% by rRT-PCR was similar to the projected PPV (89.6%.) assuming the minimum recommended sensitivity (80%) and specificity (97.0%). The NPV (85.0%) was lower compared to that computed (93.7%) assuming the minimum recommendations. Among individuals who tested negative by Ag RDT, the likelihood of being wrongly classified as uninfected was 15%. In 2021, the median positivity rate in Kenya was 7.4% (IQR, 3.2-13.1%) (37). At the observed sensitivity and specificity and the national positivity rate, PPV would range between 50.2% and 82.8%, and the NPV 92.4% - 98.2%).

The overall specificity we observed (98.5%) was high and was above the WHO recommendation for both symptomatic and asymptomatic individuals and across the other subgroups (21) and was similar to observations in similar evaluations (22,25,26,35,38–41). The Ag RDT had high specificity with less than (1.5%) of the tests (n=26) giving a false-positive result. This small proportion represents individuals who would be falsely classified as infected and unnecessarily be managed as COVID-19 cases or self-isolated and their contacts traced and quarantined.

However, we observed over half (53%) of the Ag RDT results gave false-negative results with a higher proportion (65%) among the asymptomatic individuals. This observation emphasizes the recommendation by WHO to re-test symptomatic patients with rRT-PCR when they receive a negative Ag RDT result, especially in settings where SARS-CoV-2 prevalence is ≤ 5%. Further, as a screening tool for testing contacts of confirmed cases, the 65% false negatives among asymptomatic contacts represent the proportion that would falsely be classified as virus-free, not self-isolate, and potentially infect others.

The Panbio™ Ag RDT was found to be easy to use by the majority of the users with a quick TAT for clinical decision making and implementation of preventive measures to contain transmission as observed in the literature (25,26). The main challenge we observed in conducting Ag RDT testing using the Panbio™ Ag RDT kits was dispensing the specimen onto the test device in samples collected from individuals with thick mucus. This was, however, solved by ensuring that individuals blew their noses before specimen collection.

Our field evaluation had several strengths. The evaluation was conducted in the real-world setting across multiple sites under the point-of-care conditions which had a large sample size comprising two groups (symptomatic and asymptomatic individuals) thereby representing individuals at any point of the COVID-19 disease spectrum.

### Limitation

The number of persons enrolled in the two groups, symptomatic and asymptomatic, may not reflect the true prevalence in the general population as we evaluated the Ag RDT during a period of high COVID-19 prevalence.

## Conclusion

The overall sensitivity and PPV of the Panbio™ Ag RDT were much lower than expected. Sensitivity was acceptable in symptomatic individuals with Ct value ≤30. Specificity of the Ag RDT was high and satisfactory; therefore, a positive result may not require confirmation by rRT-PCR. The kit may be useful as a rapid screening tool for only symptomatic patients in high-risk settings with limited access to RT-PCR. A negative result should be interpreted based on clinical and epidemiological information and may require retesting by rRT-PCR.

## Data Availability

All data produced in the present study are available upon reasonable request to the authors

## Acknowledgment

We acknowledge all the study participants, surveillance officers, and laboratory staff that participated in this evaluation. In addition, we thank the Ministry of Health Kenya, the Kenya Prisons Service, Washington State University (WSU) Global Health Kenya, and the County departments of Health of Kiambu, Mombasa, Nakuru, Nyeri, and Siaya for the continued support throughout the implementation of the evaluation.

## Conflict of interest

The authors declare no conflict of interest.

### Disclaimer

The findings and conclusions in this report are those of the author(s) and do not necessarily represent the official position of the U.S. Centers for Disease Control and Prevention. Use of trade names is for identification only and does not imply endorsement by [the Centers for Disease Control and Prevention/the Agency for Toxic Substances and Disease Registry], the Public Health Service, or the U.S. Department of Health and Human Services

## Supporting information

**S1 Fig:**
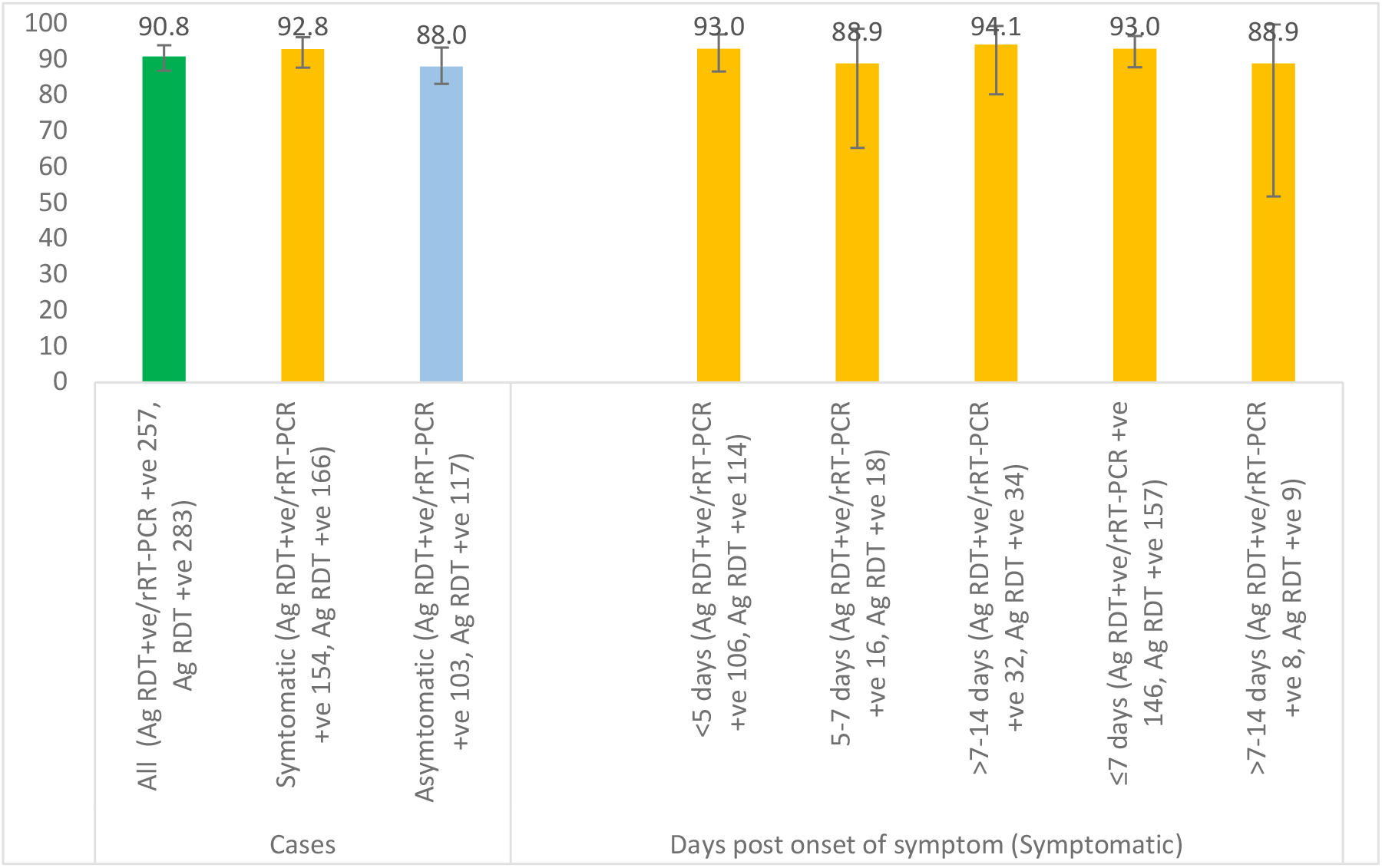
The Positive Predictive Value of the Panbio™ Ag RDT by clinical status and days post onset of symptoms, Kenya 2021

**S2 Fig:**
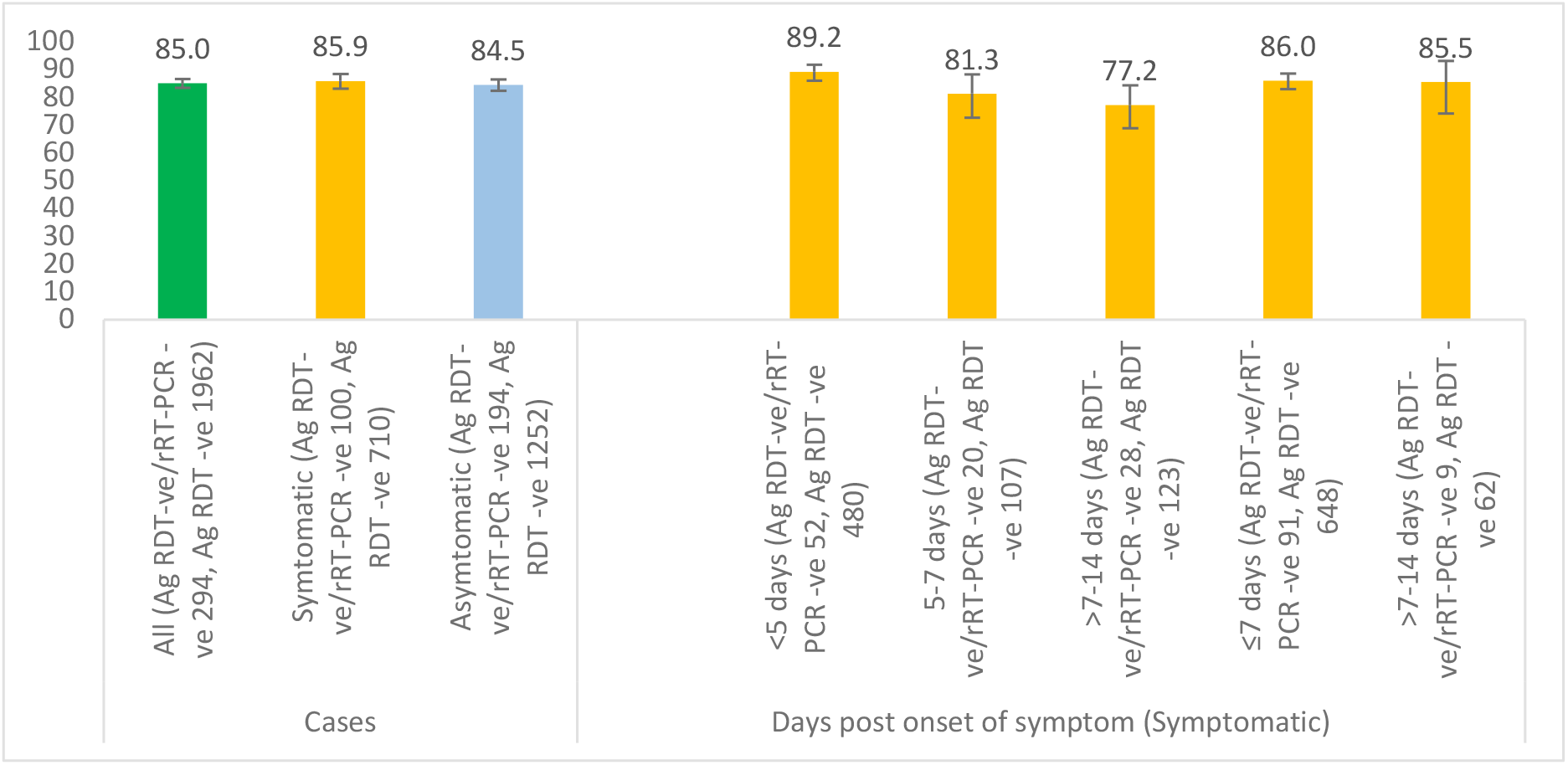
The Negative Predictive Value of the Panbio™ Ag RDT by clinical status and days post onset of symptoms, Kenya 2021

**S3 Fig:**
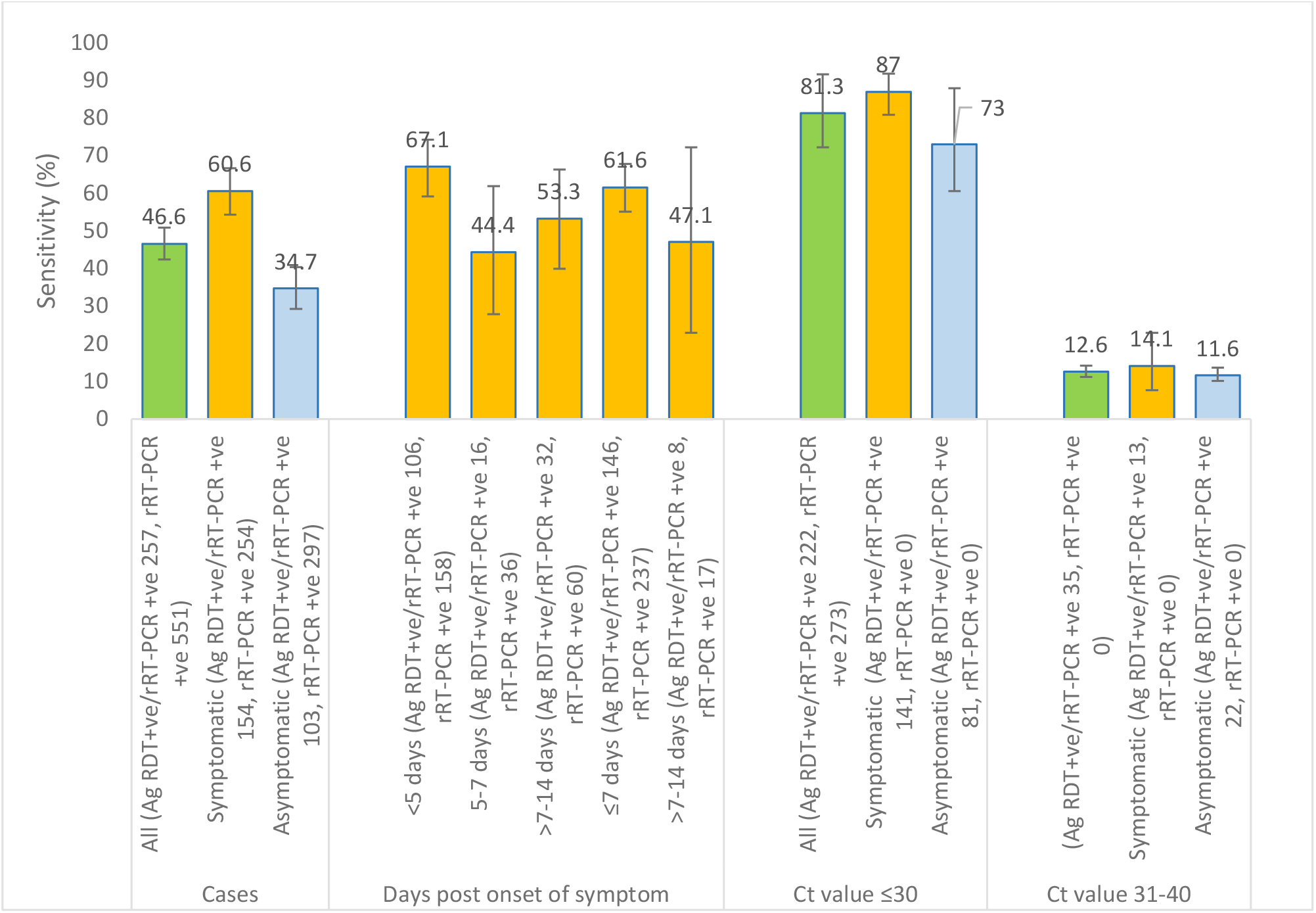
The sensitivity of the Panbio™ Ag RDT by clinical status, and Ct values

**S4 Fig:**
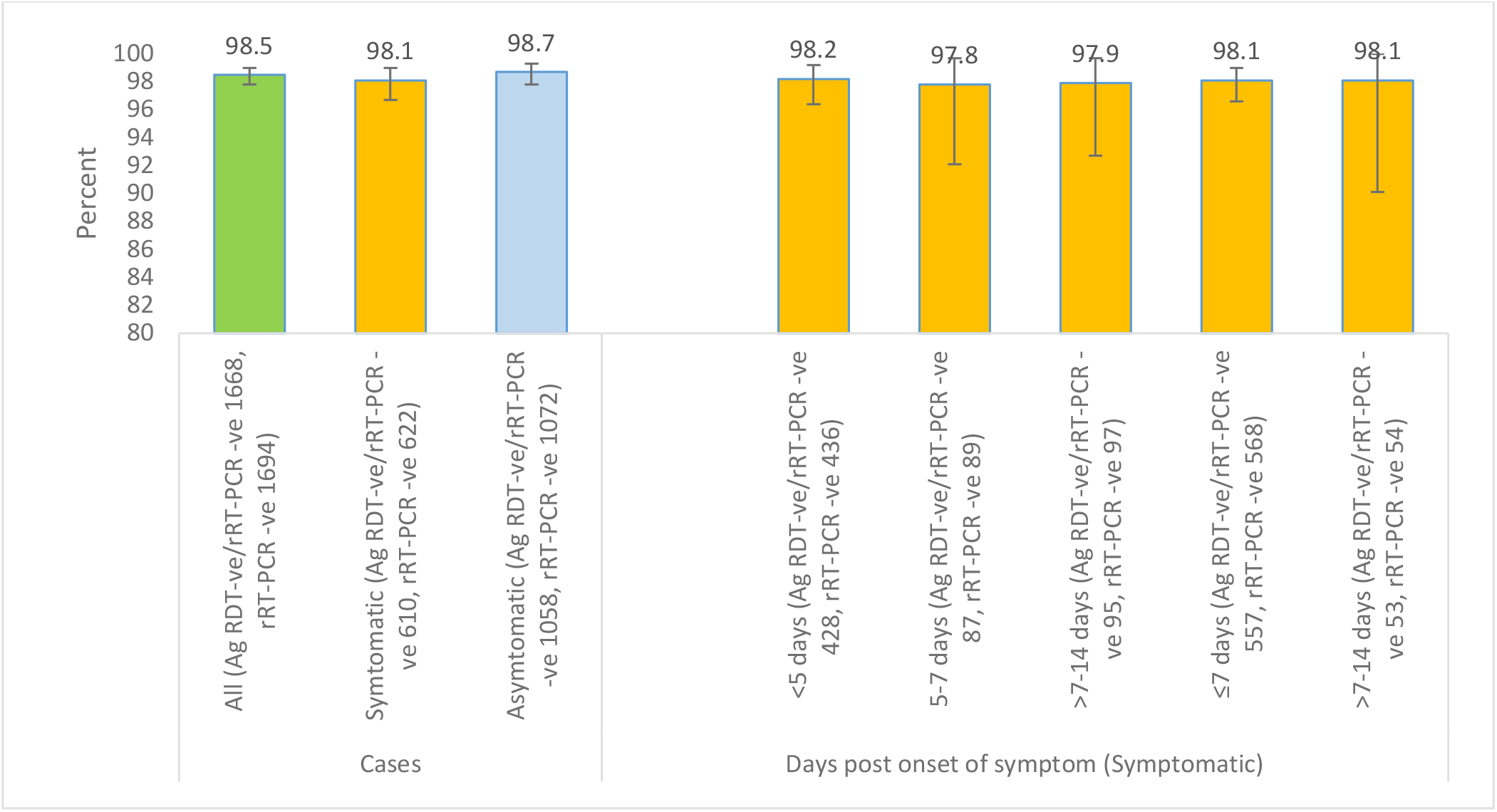
The specificity of the Panbio™ Ag RDT by clinical status and days post onset of symptoms, Kenya 2021

## References

1. Albert E, Torres I, Bueno F, Huntley D, Molla E, Fernández-Fuentes MÁ, et al. Field evaluation of a rapid antigen test (Panbio™COVID-19 Ag Rapid Test Device) for COVID-19 diagnosis in primary healthcare centres. Clin Microbiol Infect [Internet]. 2021 Mar;27(3):472.e7-472.e10. Available from: https://linkinghub.elsevier.com/retrieve/pii/S1198743X20306972

2. Barra GB, Rita THS, Mesquita PG, Jácomo RH, Nery LFA. Overcoming supply shortage for SARS-CoV-2 detection by RT-qPCR. Genes (Basel). 2021;12(1):1–10.

3. World Health Organization (WHO). Antigen-detection in the diagnosis of SARS-CoV-2 infection using rapid immunoassays Interim guidance, 11 September 2020. World Heal Organ [Internet]. 2020;(September):1–9. Available from: https://www.who.int/publications/i/item/antigen-detection-in-the-diagnosis-of-sars-cov-2infection-using-rapid-immunoassays%0Ahttps://apps.who.int/iris/handle/10665/334253

4. World Health Organization (WHO). WHO Emergency Use Listing for In vitro diagnostics (IVDs) Detecting SARS-CoV-2. Geneva [Internet]. World Health Organization (WHO). Geneva; 2020. Available from: https://extranet.who.int/pqweb/sites/default/files/documents/210217_eul_sars-cov2_product_list.pdf%0Ahttps://www.who.int/diagnostics_laboratory/201002_eul_sars_cov2_product_list.pdf?ua=1

5. Abbott. Panbio™Test Device COVID-19 Ag Rapid -In vitro diagnostic rapid test for qualitative detection of SARS-CoV-2 antigen (Ag) [Internet]. 2021. Available from: https://dam.abbott.com/en-gb/panbio/120007883-v1-Panbio-COVID-19-Ag-Nasal-AsymptomaticSe.pdf

6. Landis JR, Koch GG. The Measurement of Observer Agreement for Categorical Data. Biometrics [Internet]. 1977 Mar;33(1):159. Available from: https://www.jstor.org/stable/2529310?origin=crossref

7. Ministry of Health Kenya. Covid-19 Outbreak in Kenya Situation report 670. Vol. 670. 2021.

8. World Health Organization W. WHO Coronavirus Disease (COVID-19) Dashboard With Vaccination Data | WHO Coronavirus (COVID-19) Dashboard With Vaccination Data [Internet]. World Health Organization. 2022 [cited 2022 Feb 20]. Available from: https://covid19.who.int/region/afro/country/ke

9. Agulló V, Fernández-González M, Ortiz de la Tabla V, Gonzalo-Jiménez N, García JA, Masiá M, et al. Evaluation of the rapid antigen test Panbio COVID-19 in saliva and nasal swabs in a population-based point-of-care study. J Infect. 2021;82(5):186–230.

10. Bulilete O, Lorente P, Leiva A, Carandell E, Oliver A, Rojo E, et al. Panbio™rapid antigen test for SARS-CoV-2 has acceptable accuracy in symptomatic patients in primary health care. J Infect [Internet]. 2021 Mar;82(3):391–8. Available from: https://doi.org/10.1016/j.jinf.2021.02.014

11. Linares M, Pérez-Tanoira R, Carrero A, Romanyk J, Pérez-García F, Gómez-Herruz P, et al. Panbio antigen rapid test is reliable to diagnose SARS-CoV-2 infection in the first 7 days after the onset of symptoms. J Clin Virol [Internet]. 2020 Dec;133(January):104659. Available from: https://linkinghub.elsevier.com/retrieve/pii/S1386653220304017

12. Merino P, Guinea J, Muñoz-Gallego I, González-Donapetry P, Galán JC, Antona N, et al. Multicenter evaluation of the Panbio™COVID-19 rapid antigen-detection test for the diagnosis of SARS-CoV-2 infection. Clin Microbiol Infect. 2021;27(5):758–61.

13. Villaverde S, Domínguez-Rodríguez S, Sabrido G, Pérez-Jorge C, Plata M, Romero MP, et al. Diagnostic Accuracy of the Panbio Severe Acute Respiratory Syndrome Coronavirus 2 Antigen Rapid Test Compared with Reverse-Transcriptase Polymerase Chain Reaction Testing of Nasopharyngeal Samples in the Pediatric Population. J Pediatr [Internet]. 2021 May;232(January):287-289.e4. Available from: https://linkinghub.elsevier.com/retrieve/pii/S0022347621000342

14. Euser S, Aronson S, Manders I, van Lelyveld S, Herpers B, Sinnige J, et al. SARS-CoV-2 viral-load distribution reveals that viral loads increase with age: a retrospective cross-sectional cohort study. Int J Epidemiol [Internet]. 2022 Jan 6;50(6):1795–803. Available from: https://academic.oup.com/ije/article/50/6/1795/6366466

15. KEMRI. Genomic surveillance detects local transmission of the global variants of concern in nine counties in Kenya. 2021.

16. Kimita G, Nyataya J, Omuseni E, Sigei F, Lemtudo A, Muthanje E, et al. A genomics dissection of Kenya’s COVID-19 waves: temporal lineage replacements and dominance of imported variants of concern. 2021; Available from: https://www.researchsquare.com/article/rs-942627/v1

17. Bekliz M, Adea K, Essaidi-Laziosi M, Sacks JA, Escadafal C, Kaiser L, et al. SARS-CoV-2 rapid diagnostic tests for emerging variants. The Lancet Microbe [Internet]. 2021;2(8):e351. Available from: http://dx.doi.org/10.1016/S2666-5247(21)00147-6

18. Bekliz M, Adea K, Essaidi-Laziosi M, Sacks JA, Escadafal C, Kaiser L, et al. SARS-CoV-2 antigen-detecting rapid tests for the delta variant. The Lancet Microbe [Internet]. 2022 Feb;3(2):e90. Available from: http://dx.doi.org/10.1016/S2666-5247(21)00302-5

19. van Ogtrop ML, van de Laar TJW, Eggink D, Vanhommerig JW, van der Reijden WA. Comparison of the Performance of the PanBio COVID-19 Antigen Test in SARS-CoV-2 B.1.1.7 (Alpha) Variants versus non-B.1.1.7 Variants. Wesley Long S, editor. Microbiol Spectr [Internet]. 2021 Dec 22;9(3):1–22. Available from: https://journals.asm.org/doi/10.1128/Spectrum.00884-21

20. Jian MJ, Chung HY, Chang CK, Lin JC, Yeh KM, Chen CW, et al. SARS-CoV-2 variants with T135I nucleocapsid mutations may affect antigen test performance. Int J Infect Dis [Internet]. 2022;114:112–4. Available from: https://doi.org/10.1016/j.ijid.2021.11.006

21. World Health Organization. SARS-CoV-2 Antigen detecting rapid diagnostic test implementation projects [Internet]. 2020. p. 1–6. Available from: https://www.who.int/news-room/articles-detail/sars-cov-2-antigen-detecting-rapid-diagnostic-test-implementation-projects

22. Thirion-Romero I, Guerrero-Zúñiga DS, Arias-Mendoza DA, Cornejo-Juárez DDP, Meza-Meneses DP, Torres-Erazo DDS, et al. Evaluation of Panbio rapid antigen test for SARS-CoV-2 in symptomatic patients and their contacts: a multicenter study. Int J Infect Dis [Internet]. 2021 Dec;113:218–24. Available from: https://linkinghub.elsevier.com/retrieve/pii/S1201971221008110

23. Eleftheriou I, Dasoula F, Dimopoulou D, Lebessi E, Serafi E, Spyridis N, et al. Real-life evaluation of a COVID-19 rapid antigen detection test in hospitalized children. J Med Virol. 2021;93(10):6040–4.

24. L’Huillier AG, Lacour M, Sadiku D, Gadiri MA, De Siebenthal L, Schibler M, et al. Diagnostic accuracy of sars-cov-2 rapid antigen detection testing in symptomatic and asymptomatic children in the clinical setting. J Clin Microbiol. 2021;59(9):1–8.

25. Krüger LJ, Gaeddert M, Tobian F, Lainati F, Gottschalk C, Klein JAF, et al. The Abbott PanBio WHO emergency use listed, rapid, antigen-detecting point-of-care diagnostic test for SARS-CoV-2—Evaluation of the accuracy and ease-of-use. Mantis NJ, editor. PLoS One [Internet]. 2021 May 27;16(5):e0247918. Available from: https://dx.plos.org/10.1371/journal.pone.0247918

26. FIND. FIND Evaluation of Abbott Panbio COVID-19 Ag Rapid Test Device External Report. 2020;(December):1–4. Available from: https://www.finddx.org/wp-content/uploads/2020/12/Panbio_Ag-Public-Report_v2.1.pdf

27. Alemany A, Baró B, Ouchi D, Rodó P, Ubals M, Corbacho-Monné M, et al. Analytical and clinical performance of the panbio COVID-19 antigen-detecting rapid diagnostic test. J Infect [Internet]. 2021 May;82(5):186–230. Available from: https://linkinghub.elsevier.com/retrieve/pii/S0163445321000050

28. Almadhi MA, Abdulrahman A, Sharaf SA, AlSaad D, Stevenson NJ, Atkin SL, et al. The high prevalence of asymptomatic SARS-CoV-2 infection reveals the silent spread of COVID-19. Int J Infect Dis [Internet]. 2021;105:656–61. Available from: https://doi.org/10.1016/j.ijid.2021.02.100

29. Strutner J, Ramchandar N, Dubey S, Gamboa M, Vanderpool MK, Mueller T, et al. Comparison of Reverse-Transcription Polymerase Chain Reaction Cycle Threshold Values From Respiratory Specimens in Symptomatic and Asymptomatic Children With Severe Acute Respiratory Syndrome Coronavirus 2 Infection. Clin Infect Dis [Internet]. 2021 Nov 16;73(10):1790–4. Available from: https://academic.oup.com/cid/article/73/10/1790/6266745

30. Zuin M, Gentili V, Cervellati C, Rizzo R, Zuliani G. Viral load difference between symptomatic and asymptomatic COVID-19 patients: Systematic review and meta-analysis. Infect Dis Rep. 2021;13(3):645–53.

31. Lavezzo E, Franchin E, Ciavarella C, Cuomo-Dannenburg G, Barzon L, Del Vecchio C, et al. Suppression of a SARS-CoV-2 outbreak in the Italian municipality of Vo’. Nature [Internet]. 2020 Aug 20;584(7821):425–9. Available from: http://www.nature.com/articles/s41586-020-2488-1

32. Cardillo L, de Martinis C, Viscardi M, Esposito C, Sannino E, Lucibelli G, et al. SARS-CoV-2 quantitative real time PCR and viral loads analysis among asymptomatic and symptomatic patients: an observational study on an outbreak in two nursing facilities in Campania Region (Southern Italy). Infect Agent Cancer. 2021;16(1):1–7.

33. Hasanoglu I, Korukluoglu G, Asilturk D, Cosgun Y, Kalem AK, Altas AB, et al. Higher viral loads in asymptomatic COVID-19 patients might be the invisible part of the iceberg. Infection [Internet]. 2021;49(1):117–26. Available from: https://doi.org/10.1007/s15010-020-01548-8

34. He X, Lau EHY, Wu P, Deng X, Wang J, Hao X, et al. Temporal dynamics in viral shedding and transmissibility of COVID-19. Nat Med. 2020;26(5):672–5.

35. Berger A, Nsoga MTN, Perez-Rodriguez FJ, Aad YA, Sattonnet-Roche P, Gayet-Ageron A, et al. Diagnostic accuracy of two commercial SARSCoV-2 antigen-detecting rapid tests at the point of care in community-based testing centers. PLoS One. 2021;16(3 March 2021):1–12.

36. Ferté T, Ramel V, Cazanave C, Lafon M-E, Bébéar C, Malvy D, et al. Accuracy of COVID-19 rapid antigenic tests compared to RT-PCR in a student population: The StudyCov study. J Clin Virol [Internet]. 2021 Aug;141(January):104878. Available from: https://linkinghub.elsevier.com/retrieve/pii/S1386653221001451

37. Hasell J, Mathieu E, Beltekian D, Macdonald B, Giattino C, Ortiz-Ospina E, et al. A cross-country database of COVID-19 testing. Sci Data [Internet]. 2020 Dec 8 [cited 2022 Feb 20];7(1):345. Available from: https://ourworldindata.org/coronavirus-testing

38. Albert E, Torres I, Bueno F, Huntley D, Molla E, Fernández-Fuentes MÁ, et al. Field evaluation of a rapid antigen test (Panbio™COVID-19 Ag Rapid Test Device) for COVID-19 diagnosis in primary healthcare centres. Clin Microbiol Infect [Internet]. 2021;27(3):472.e7-472.e10. Available from: https://doi.org/10.1016/j.cmi.2020.11.004

39. Christensen A, Larsdatter M, Landaas E, Bragstad K, Kran A-MB, Tollånes M, et al. Evaluation of Abbot’s Panbio COVID-19 rapid antigen test in Norway [Internet]. 2020. Available from: https://www.helsedirektoratet.no/rapporter/evaluation-of-abbots-panbio-covid-19-rapid-antigen-test-in-norway/EvaluationofAbbotsPanbioOVID-1BQZKqdp2CV3QV5nUEsqSg1ygegLmqRygj.pdf/_/attachment/inline/b3306b98-c0e0-4e96-aa62-3ca5a99f5367:10fe6f072721ece7aee

40. Torres I, Poujois S, Albert E, Colomina J, Navarro D. Evaluation of a rapid antigen test (Panbio™COVID-19 Ag rapid test device) for SARS-CoV-2 detection in asymptomatic close contacts of COVID-19 patients. Clin Microbiol Infect [Internet]. 2021 Apr;27(4):636.e1-636.e4. Available from: https://linkinghub.elsevier.com/retrieve/pii/S1198743X20307825

41. Dinnes J, Deeks JJ, Berhane S, Taylor M, Adriano A, Davenport C, et al. Rapid, point-of-care antigen and molecular-based tests for diagnosis of SARS-CoV-2 infection. Cochrane Database Syst Rev [Internet]. 2021 Mar 24;2021(4). Available from: http://doi.wiley.com/10.1002/14651858.CD013705.pub2

